# Incidence of Symptoms Associated with Post-Acute Sequelae of SARS-CoV-2 infection in Non-Hospitalized Vaccinated Patients Receiving Nirmatrelvir-Ritonavir

**DOI:** 10.1101/2023.04.05.23288196

**Authors:** Rushin Patel, Sourbha Dani, Sumanth Khadke, Javaria Ahmad, Jui Shah, Neev Mehta, Kenneth Wener, Daniel P McQuillen, George Abraham, Jeremy Faust, Jason Maley, Smita Patel, Janet Mullington, Robert M. Wachter, Anne Mosenthal, Paul E. Sax, Sarju Ganatra

## Abstract

**Background:** The role of Nirmatrelvir plus ritonavir (NMV-r) in preventing post-acute sequelae of SARS-CoV-2 infection (PASC) is unknown. The objective of this study is to assess the effect of NMV-r in non-hospitalized, vaccinated patients on the occurrence of PASC.

**Methods:** We performed a comparative retrospective cohort study utilizing data from the TriNetX research network, including vaccinated patients ≥18 years old who subsequently developed Covid-19 between December 2021-April 2022. Cohorts were based on NMV-r administration within five days of diagnosis. Based on previously validated broad and narrow definitions, the main outcome was the presence of symptoms associated with PASC. Outcomes were assessed between 30-180 days and 90-180 days after the index Covid-19 infection.

**Results:** 1,004 patients remained in each cohort after propensity-score matching. PASC (broad definition) occurred in 425 patients (42%) in the NMV-r cohort, vs. 480 patients (48%) in the control cohort (OR 0.8 CI 0.67-0.96; p=0.01) from 30-180 days and in 273 patients (27%) in the NMV-r cohort, as compared to 347 patients (35%) in the control cohort (OR 0.707, CI 0.59-0.86; p<0.001) from 90-180 days. Narrowly defined PASC was reported in 337 (34%) patients in the NMV-r and 404 (40%) in the control cohort between 30-180 days (OR=0.75, CI 0.62-0.9, p=0.002) and in 221 (22%) in the NMV-r cohort as compared to in 278 (28%) patients in the control cohort (OR=0.7, CI 0.63-0.9, p=0.003) between 90 -180 days.

**Conclusions:** NMV-r treatment in non-hospitalized vaccinated patients with Covid-19 was associated with a reduction in the development of symptoms commonly observed with PASC and healthcare utilization.

**Key Points:** Assessment of Nirmatrelvir plus ritonavir (NMV-r) in preventing post-acute sequelae of SARS-CoV-2 infection (PASC), based on broad and narrow definitions in non-hospitalized, vaccinated patients between 30-180 days and 90-180 days.

## Introduction

### Background

Coronavirus Disease 2019 (Covid-19) caused by the SARS-CoV-2 virus can present with varied clinical presentations ranging from an asymptomatic or mildly symptomatic disease that can be managed in the outpatient setting to severe illness requiring hospitalization. In addition to acute illness, Covid-19 can result in a lingering long-term cluster of symptoms lasting for months [1]. Numerous terms, including “long Covid,” “post-Covid condition,” and post-acute sequelae of SARS-CoV2 infection (PASC), have been used to describe these syndromes. According to the World Health Organization, as of August 2022, 581 million people worldwide have recovered from Covid-19 [2]. The reported prevalence of PASC is 6% to 54% [3, 4], with this wide variation due to variable definitions of PASC and different populations studied. Even with the lower prevalence estimate, these numbers suggest a staggering global disease burden, making it a condition with enormous medical, public health, and economic consequences.

Advances in vaccination and therapies, along with the impact of widespread immunity acquired from prior infection, have markedly lowered the case-fatality rate of Covid-19. In addition, a significant advance has been the availability of effective outpatient therapeutics, including nirmatrelvir plus ritonavir (NMV-r) [5], molnupiravir [6], remdesivir [7], and various monoclonal antibodies [8].□ However, while the efficacy of these treatments in lowering the risk of acute Covid has been well established, there are currently no published data in peer-reviewed literature regarding their effectiveness in reducing the frequency of PASC.

Of the available options for outpatient management of Covid-19, NMV-r is a favored treatment based on the results of a pivotal placebo-controlled trial in unvaccinated patients with the mild-moderate disease [5]. In this study, NMV-r resulted in an 89% reduction in risk of hospitalization or all-cause mortality compared to the placebo, leading to the Emergency Use Authorization of the drug [5]. Additionally, in several observational studies, NMV-r treatment was associated with reduced all-cause mortality, emergency room (ER) visits, and hospitalizations in broader patient groups, including high-risk vaccinated Covid-19 patients (45% relative risk reduction) [9].

### Objective

Given the frequency and consequences of PASC, determining whether antiviral therapy is effective in preventing PASC is of great importance. While ideally, these data would come from prospective randomized placebo-controlled clinical trials; such studies have not yet reported these outcomes with sufficient follow-up to answer this question. To address this data gap, we utilized the electronic health records (EHR)-based, curated real-world data of the TriNetX research network[10] to describe the frequency of PASC-associated symptoms among those treated versus not treated with NMV-r.

## Methods

### Study Oversight

Data analysis and review of manuscripts were performed by all the above authors. Institutional review board (IRB) approval was exempted from Lahey Clinic IRB as deidentified data was obtained from a research network database. The study findings are reported per the Strengthening the Reporting of Observational Studies in Epidemiology (STROBE) guidelines for cohort studies.

### Data Source and Study Setting

This study utilized the TriNetX Analytics Network – Research Network, a multicenter federated health research network using de-identified data from electronic health records (EHRs) from participating healthcare organizations, including academic medical centers and specialty physician practices, and community hospitals. The research network contains data on more than 88 million patients. While the data are in aggregate de-identified form, the built-in analytics allows for the generation of patient-level data for cohort selection and matching, analyzing incidence and prevalence of events in a cohort, and comparing characteristics and outcomes between matched cohorts. More information on the database can be found online [10].

### Study Population and Design

The TriNetX research network was searched, and data curation was performed on October 17, 2022. A comparative retrospective cohort study was conducted, which included non-hospitalized patients ≥18 years who were vaccinated and subsequently developed Covid-19 between December 1, 2021, and April 17, 2022, at least one month after vaccination. Key exclusion criteria were treatment with a monoclonal antibody, convalescent plasma, or molnupiravir for the index case of Covid-19 and hospitalization for Covid-19. Patients were further categorized based on whether they were prescribed NMV-r within five days of diagnosis.

Patients with PASC were identified using two definitions based on the ICD code. Validated diagnostic, procedure and laboratory codes were utilized to define the vaccination status and Covid-19 diagnosis. Identification of patients who were prescribed NMV-r was completed using the National Library of Medicine RxNorm terminology.

Cohorts were matched using propensity score matching (PSM). Outcomes were analyzed from 30 to 180 days and between 90 to 180 days after the index diagnosis of Covid-19.

The Supplementary Appendix provides additional inclusion and exclusion criteria and information.

### Study Endpoints/variables

#### Main Composite Endpoint

Since no validated diagnostic tests exist, PASC is defined by patient-reported symptoms. To account for the lack of a standard definition and the wide range of estimates on the prevalence of PASC after Covid-19, we defined PASC both broadly and narrowly, using prior published reports [11, 12].

1) Broadly-defined PASC: included a list of the constitutional, cardio-respiratory, gastrointestinal, musculoskeletal, and nervous system, and/or mood/cognitive disorder symptoms [11].

2) Narrowly-defined PASC: includes fatigue with myalgia or mood disorder symptoms, cognitive disorder, or respiratory symptoms [12].

Other outcomes, including complications and healthcare utilization, were also defined using validated codes.

In addition to the absence of a specific diagnostic test, there is a lack of agreement regarding how long symptoms persist after Covid-19 qualify as PASC. Therefore, the outcomes were analyzed beyond the first 30 days, beyond the first 90 days, and up to a total of 180 days following the index diagnosis of Covid-19.

#### Other Endpoints

Secondary endpoints included individual symptoms of PASC. We also assessed healthcare utilization, including diagnostic imaging and cardiovascular testing.

### Statistical Analysis

Non-hospitalized vaccinated patients who developed Covid-19 at least four weeks after vaccination were divided into two cohorts based on their use of NMV-r within five days of diagnosis. These two cohorts (NMV-r and non-NMV-r cohorts) were compared using independent sample t-tests for continuous variables, which are reported as mean (range). Categorical variables are reported as counts (%) and compared using the Chi-square (χ2) test. To control for baseline differences in the patient cohorts, we performed 1:1 Propensity Score matching (PSM) for characteristics of clinical relevance utilizing a built-in algorithm that uses the greedy nearest-neighbor algorithm with a caliper of 0.1 pooled standard deviations. First, symptoms of PASC seen between 30 to 180 days after developing Covid-19 were assessed. Subsequently, symptoms of PASC seen between 90 to 180 days were also assessed. Any characteristic with a standardized mean difference between cohorts lower than 0.1 was considered well-matched. After propensity matching, odds ratios with 95% confidence intervals were calculated for primary and secondary outcomes using the χ2 for the measures of association. Relative risk reduction was calculated as the division of the absolute risk reduction between the treatment (NMV-r) and control (non-NMV-r) cohorts by the absolute risk of the control group. Statistical analyses were completed using the TriNetX online platform using R for statistical computing.

## Results

### Study Population

279,380 vaccinated individuals who tested positive for Covid-19 and were not hospitalized during the study were identified. Of these, 1,004 were treated with NMV-r within five□days of diagnosis and had sufficient follow-up for inclusion, and 278,376 were not treated with NVM-r. After propensity score matching, 1,004 patients were included in each cohort (Figure 1).

**Figure 1:**
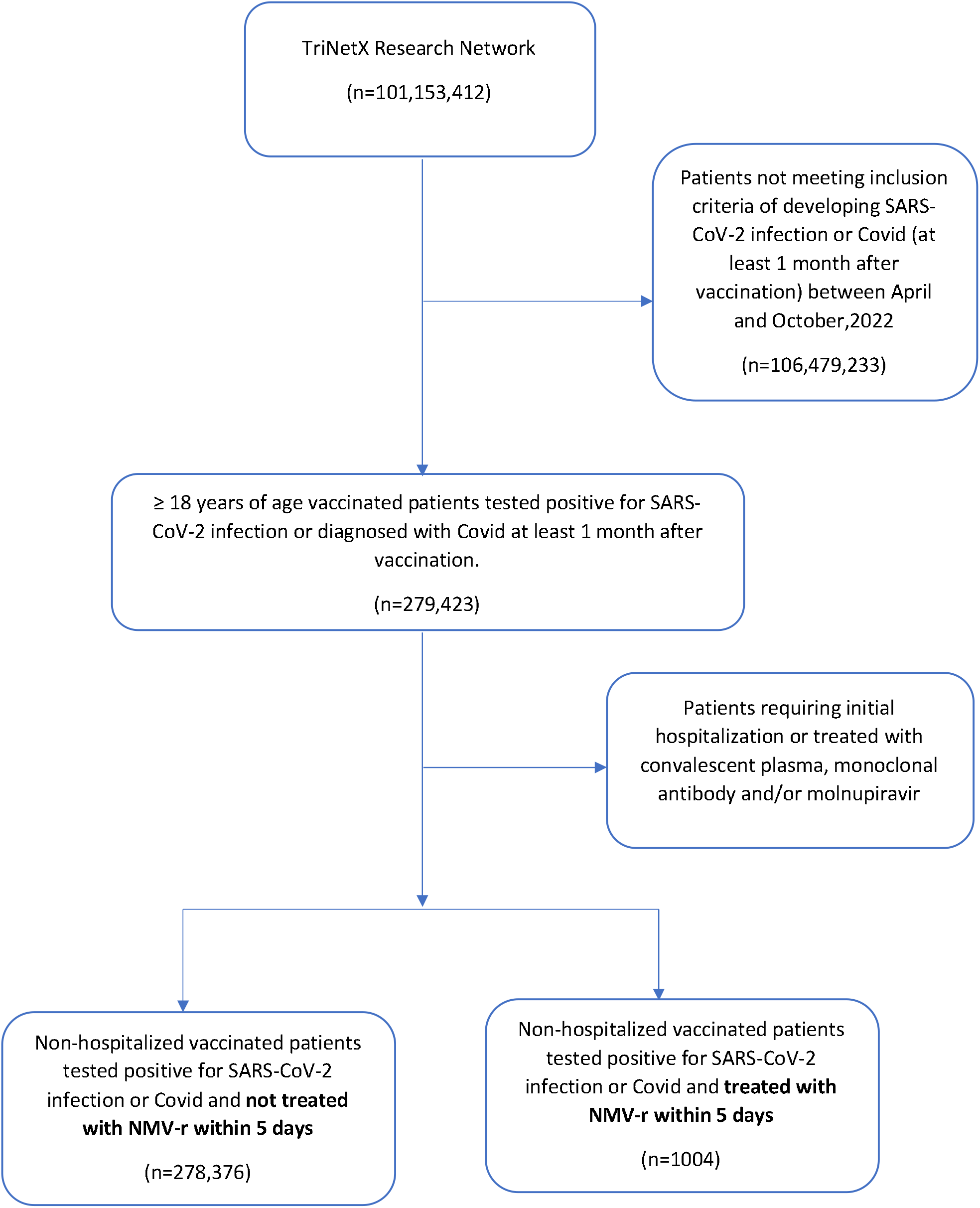
CONSORT diagram. This figure illustrates the proportion of vaccinated, non-hospitalized patients who tested positive for SARS-CoV-2 infection or were diagnosed with Covid-19 stratified by the use of NMV-r. Abbreviations: CONSORT, Consolidated Standards of Reporting Trials; Covid-19, coronavirus disease 2019; HCO, healthcare organization; NMV-r, nirmatrelvir plus ritonavir; SARS-CoV-2, severe acute respiratory syndrome coronavirus 2.

**Figure 2.**
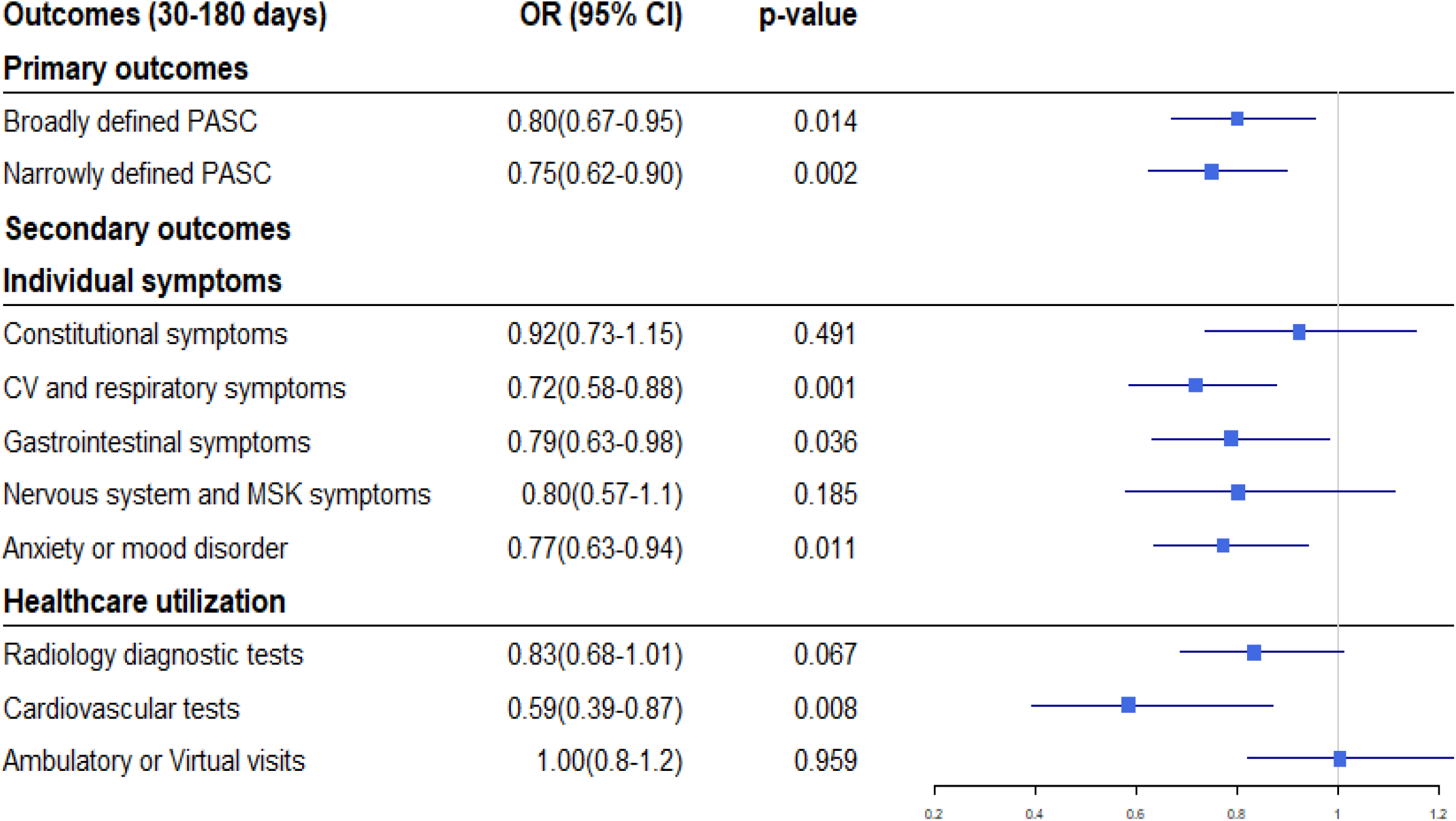
Forest Plot (30-180 Days) This forest plot depicts the primary and secondary outcomes within 30-180 days after the diagnosis of Covid-19. Abbreviations, PASC: post-acute sequelae of SARS-CoV-2 infection; CV: Cardiovascular; MSK: Musculoskeletal

**Figure 3.**
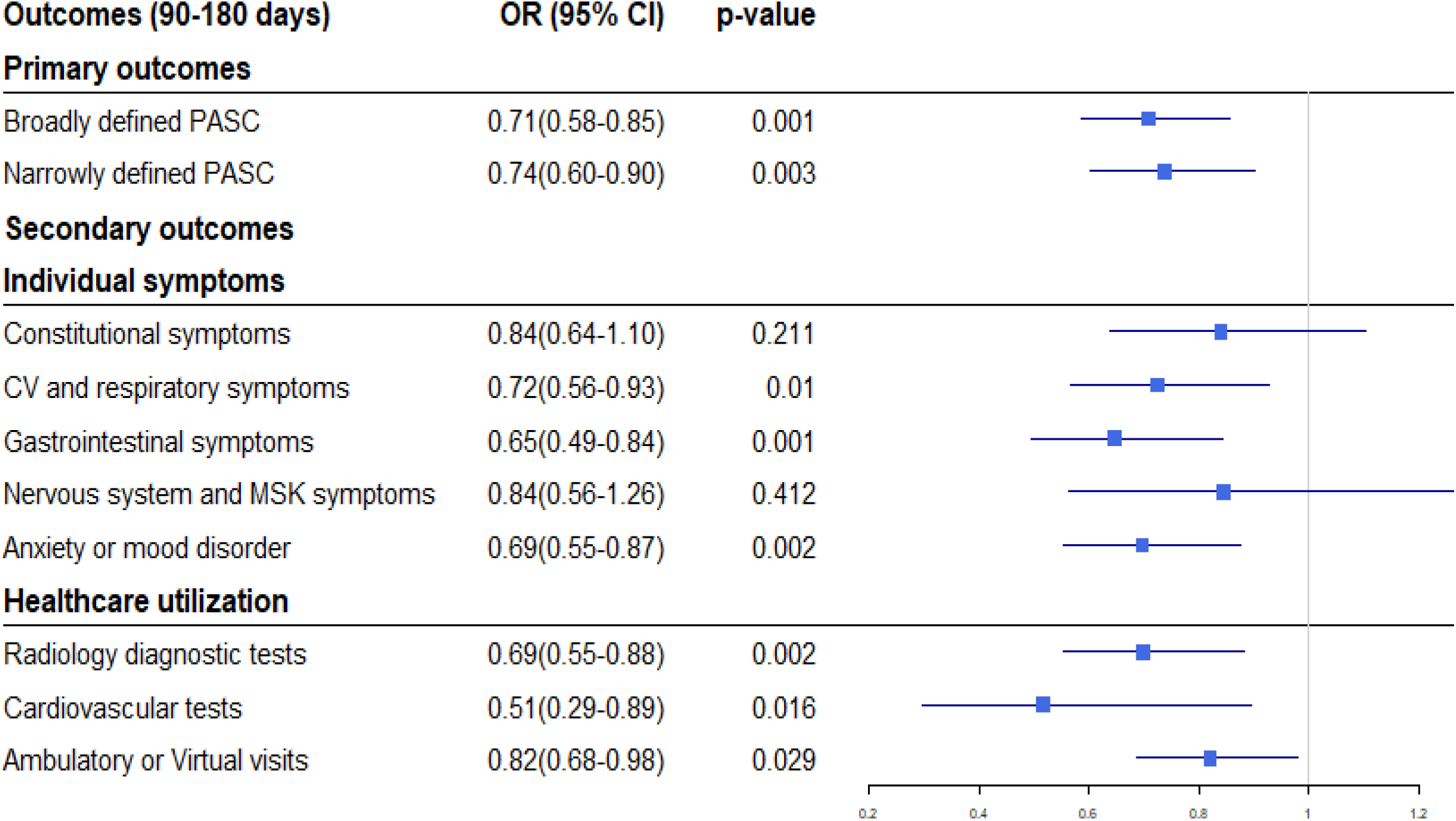
Forest Plot (90-180 Days) This forest plot depicts the primary and secondary outcomes within 90-180 days after the diagnosis of Covid-19. Abbreviations, PASC: post-acute sequelae of SARS-CoV-2 infection; CV: Cardiovascular; MSK: Musculoskeletal

### Patient Demographics

Table 1 outlines the baseline characteristics of each cohort before and after propensity matching. Before propensity matching, the mean age of patients treated with NMV-r was higher (mean age 57.2 ±16.7 versus 49.7 ± 18.0; SD 0.430), and there were racial differences - the proportion of the Caucasian population was higher in the NMV-r cohort compared to non-NMV-r cohort (82.0% in NMV-r vs. 69.7%). In contrast, the proportion of African Americans was higher in the non-NMV-r cohort (17.1% in non-NMV-r vs. 7.9% in NMV-r). In addition, patients in the NMV-r cohort had a higher prevalence of cardiovascular risk factors (hypertension, hyperlipidemia, diabetes mellitus). Compared to the comparison cohort, more patients were on medications for cardiovascular disease (beta-blockers, diuretics, ACE inhibitors, ARBs, aspirin, statins). In the NMV-r cohort, the prevalence was also higher for neoplasms, systemic connective tissue disorders, and chronic lower respiratory disease. □All baseline characteristics between the two cohorts, including healthcare utilization, were propensity matched, with no residual difference (standard difference for all included covariates was <0.1).

**Table 1.** Baseline Characteristics

|  | Before Propensity matching |  |  | After Propensity Matching |  |  |
| --- | --- | --- | --- | --- | --- | --- |
| Baseline Characteristics | Received NMVr (n=1004) | No NMVr (n=278,376) | SD | Received NMVr (n=1004) | No NMVr (n=1004) | SD |
| Demographics |  |  |  |  |  |  |
| Mean Age | 57.2 +/- 16.7 | 49.7 +/- 18.0 | 0.430 | 57.2+/-16.7 | 56.7 +/- 16.6 | 0.028 |
| Female | 644 (64.1%) | 172,955 (62.1%) | 0.042 | 644 (64.1%) | 661 (65.8%) | 0.036 |
| Caucasian | 823 (82.0%) | 194,158 (69.7%) | 0.289 | 823 (82.0%) | 801 (79.8%) | 0.056 |
| African American | 79 (7.9%) | 47,520 (17.1%) | 0.281 | 79 (7.9%) | 84 (8.4%) | 0.018 |
| Non-Hispanic/Latino | 795 (79.2%) | 166,061 (59.7%) | 0.434 | 795 (79.2%) | 794 (79.1%) | 0.002 |
| Prior Healthcare Utilization |  |  |  |  |  |  |
| Ambulatory Visit | 979 (97.5%) | 219,296 (78.8%) | 0.605 | 979 (97.5%) | 979 (97.5%) | <0.001 |
| ER Visit | 484 (48.2%) | 129,143 (46.4%) | 0.036 | 484 (48.2%) | 500 (49.8%) | 0.032 |
| Hospital Inpatient Services | 204 (20.3%) | 45,112 (16.2%) | 0.107 | 204 (20.3%) | 199 (19.8%) | 0.012 |
| Hospital Observation Services | 109 (10.9%) | 24,021 (8.6%) | 0.075 | 109 (10.9%) | 116 (11.6%) | 0.022 |
| Critical Care Services | 46 (4.6%) | 12,413 (4.5%) | 0.006 | 46 (4.6%) | 45 (4.5%) | 0.005 |
| Comorbidities |  |  |  |  |  |  |
| Hypertension | 573 (57.1%) | 112,181 (40.3%) | 0.340 | 573 (57.1%) | 587 (58.5%) | 0.028 |
| Hyperlipidemia | 547 (54.5%) | 102,803 (36.9%) | 0.358 | 547 (54.5%) | 524 (52.2%) | 0.046 |
| Diabetes Mellitus | 243 (24.2%) | 55,360 (19.9%) | 0.104 | 243 (24.2%) | 239 (23.8%) | 0.009 |
| Chronic Lower Respiratory Disease | 332 (33.1%) | 66,278 (23.8%) | 0.206 | 332 (33.1%) | 343 (34.2%) | 0.023 |
| Chronic Kidney Disease | 110 (11.0%) | 29,014 (10.4%) | 0.017 | 110 (11.0%) | 101 (10.1%) | 0.029 |
| Atrial Fibrillation/ Flutter | 49 (4.9%) | 16,769 (6.0%) | 0.050 | 49 (4.9%) | 50 (5.0%) | 0.005 |
| Ischemic Heart Disease | 192 (19.1%) | 37,046 (13.3%) | 0.158 | 192 (19.1%) | 190 (18.9%) | 0.005 |
| Heart Failure | 56 (5.6%) | 19,606 (7.0%) | 0.060 | 56 (5.6%) | 54 (5.4%) | 0.009 |
| Ischemic Stroke | 56 (5.6%) | 13,684 (4.9%) | 0.030 | 56 (5.6%) | 52 (5.2%) | 0.018 |
| Malignancy | 463 (46.1%) | 91,001 (32.7%) | 0.277 | 463 (46.1%) | 436 (43.4%) | 0.054 |
| Demyelinating Disease | 22 (2.2%) | 2,376 (0.9%) | 0.109 | 22 (2.2%) | 11 (1.1%) | 0.086 |
| Systematic Connective Tissue Disorder | 90 (9.0%) | 12,018 (4.3%) | 0.187 | 90 (9.0%) | 70 (7.0%) | 0.074 |
| BMI > 30 | 31.6 +/- 7.2 | 30.4 +/- 7.2 | 0.177 | 31.6 +/- 7.2 | 31.3 +/- 7.8 | 0.043 |
| Medications |  |  |  |  |  |  |
| Beta Blockers | 366 (36.5%) | 74,596 (26.8%) | 0.209 | 366 (36.5%) | 372 (37.1%) | 0.012 |
| Diuretics | 348 (34.7%) | 69,406 (24.9%) | 0.214 | 348 (34.7%) | 344 (34.3%) | 0.008 |
| ACE inhibitors | 213 (21.2%) | 46,596 (16.7%) | 0.114 | 213 (21.2%) | 198 (19.7%) | 0.037 |
| Angiotensin Receptor Blocker | 201 (20.0%) | 36,065 (13.0%) | 0.191 | 201 (20.0%) | 204 (20.3%) | 0.007 |
| Aspirin | 296 (29.5%) | 59,906 (21.5%) | 0.183 | 296 (29.5%) | 298 (29.7%) | 0.004 |
| Anticoagulants | 285 (28.4%) | 67,466 (24.2%) | 0.094 | 285 (28.4%) | 274 (27.3%) | 0.024 |
| Statin | 388 (38.6%) | 78,077 (28.0%) | 0.226 | 388 (38.6%) | 374 (37.3%) | 0.029 |
| Immune Suppressants | 54 (5.4%) | 12,390 (4.5%) | 0.043 | 54 (5.4%) | 47 (4.7%) | 0.032 |
| Antineoplastics | 141 (14.0%) | 27,247 (9.8%) | 0.132 | 141 (14.0%) | 139 (13.8%) | 0.006 |
| Antidepressant | 427 (42.5%) | 95,556 (34.3%) | 0.169 | 427 (42.5%) | 469 (46.7%) | 0.084 |
| **Laboratory Tests** |  |  |  |  |  |  |
| Hemoglobin (g/dL) | 13.6 +/- 1.6 | 13.3 +/- 1.9 | 0.178 | 13.6 +/- 1.6 | 13.4 +/- 1.8 | 0.100 |
| Platelets (10<sup>3</sup>/uL) | 252.0 +/- 75.7 | 255.6 +/- 78.5 | 0.047 | 252.0 +/- 75.7 | 255.9 +/- 75.5 | 0.052 |
| Creatinine (mg/dL) | 0.9 +/- 0.3 | 1.0 +/- 2.4 | 0.074 | 0.9 +/- 0.3 | 0.9 +/- 0.6 | 0.081 |
| Total Cholesterol (mg/dL) | 180.0 +/- 40.3 | 178.2 +/- 43.5 | 0.042 | 180.0 +/- 40.3 | 178.0 +/- 44.7 | 0.046 |
| LDL (mg/dL) | 101.3 +/- 34.0 | 102.0 +/- 35.8 | 0.021 | 101.3 +/- 34.0 | 102.3 +/- 36.2 | 0.029 |
| Triglyceride (mg/dL) | 129.0 +/- 83.3 | 120.8 +/- 105.2 | 0.086 | 129.0 +/- 83.3 | 128.0 +/- 88.8 | 0.012 |
| Hemoglobin A1c | 6.0 +/- 1.3 | 6.2 +/- 1.7 | 0.123 | 6.0 +/- 1.3 | 6.0 +/- 1.4 | 0.023 |
| Left Ventricular Ejection Fraction (LVEF) (%) | 60.0 +/- 11.2 | 57.4 +/- 13.0 | 0.218 | 60.0 +/- 11.2 | 60.0 +/- 11.0 | 0.004 |

Abbreviations: ACE – Angiotensin Converting Enzyme, BMI – Body Mass Index, LDL – Low-Density Lipoprotein, SD – Standard Difference

**Table 2:**
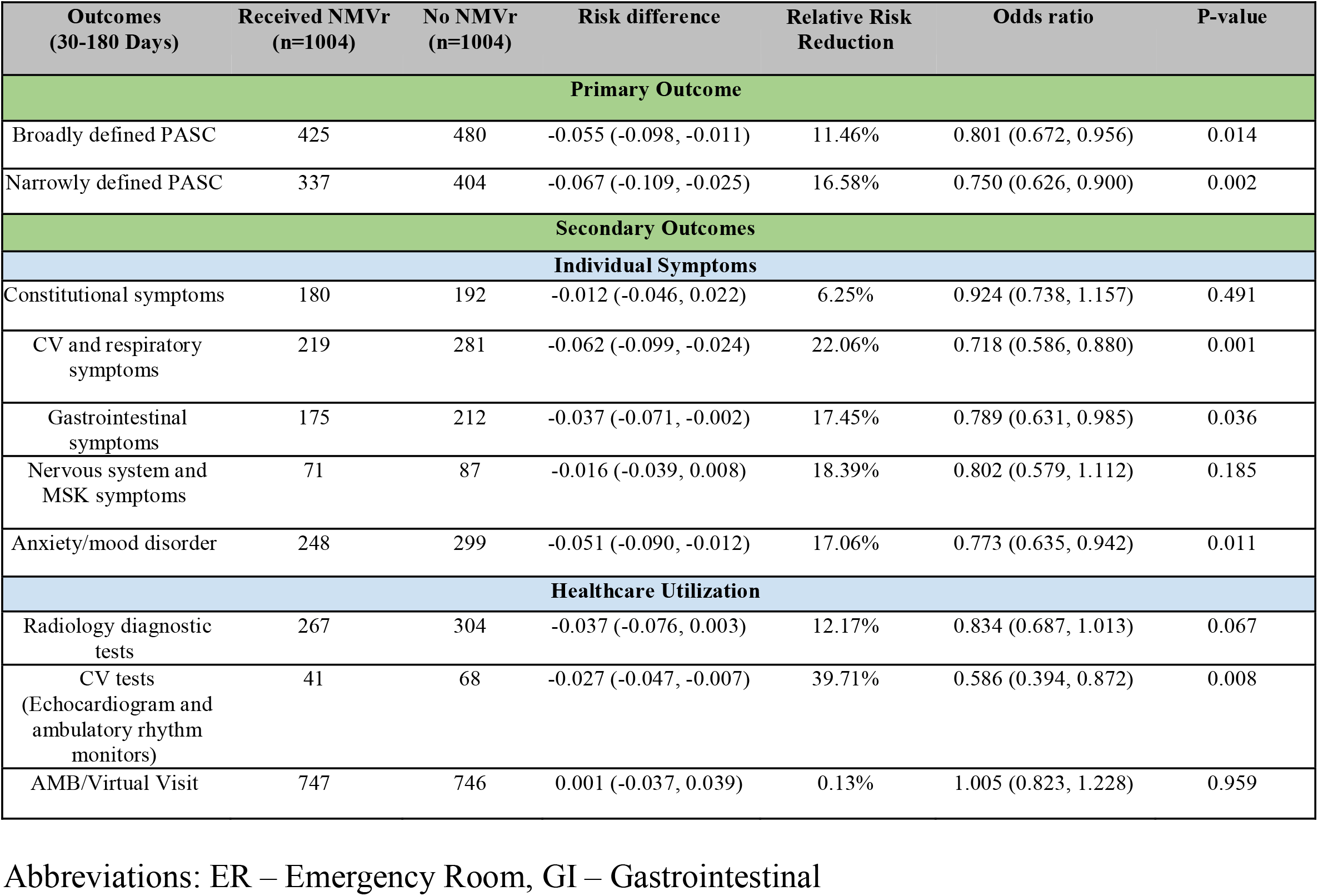
PASC Outcomes 30-180 Days

| Outcomes<br>(30-180 Days) | Received NMVr<br>(n=1004) | No NMVr<br>(n=1004) | Risk difference | Relative Risk<br>Reduction | Odds ratio | P-value |
| --- | --- | --- | --- | --- | --- | --- |
| **Primary Outcome** |  |  |  |  |  |  |
| Broadly defined PASC | 425 | 480 | -0.055 (-0.098, -0.011) | 11.46% | 0.801 (0.672, 0.956) | 0.014 |
| Narrowly defined PASC | 337 | 404 | -0.067 (-0.109, -0.025) | 16.58% | 0.750 (0.626, 0.900) | 0.002 |
| **Secondary Outcomes** |  |  |  |  |  |  |
| **Individual Symptoms** |  |  |  |  |  |  |
| Constitutional symptoms | 180 | 192 | -0.012 (-0.046, 0.022) | 6.25% | 0.924 (0.738, 1.157) | 0.491 |
| CV and respiratory<br>symptoms | 219 | 281 | -0.062 (-0.099, -0.024) | 22.06% | 0.718 (0.586, 0.880) | 0.001 |
| Gastrointestinal<br>symptoms | 175 | 212 | -0.037 (-0.071, -0.002) | 17.45% | 0.789 (0.631, 0.985) | 0.036 |
| Nervous system and<br>MSK symptoms | 71 | 87 | -0.016 (-0.039, 0.008) | 18.39% | 0.802 (0.579, 1.112) | 0.185 |
| Anxiety/mood disorder | 248 | 299 | -0.051 (-0.090, -0.012) | 17.06% | 0.773 (0.635, 0.942) | 0.011 |
| **Healthcare Utilization** |  |  |  |  |  |  |
| Radiology diagnostic<br>tests | 267 | 304 | -0.037 (-0.076, 0.003) | 12.17% | 0.834 (0.687, 1.013) | 0.067 |
| CV tests<br>(Echocardiogram and<br>ambulatory rhythm<br>monitors) | 41 | 68 | -0.027 (-0.047, -0.007) | 39.71% | 0.586 (0.394, 0.872) | 0.008 |
| AMB/Virtual Visit | 747 | 746 | 0.001 (-0.037, 0.039) | 0.13% | 1.005 (0.823, 1.228) | 0.959 |

Abbreviations: ER – Emergency Room, GI – Gastrointestinal

**TABLE 3.** PASC Outcomes 90-180 Days

| Outcomes<br>(90-180 days) | Received NMVr<br>(n=1004) | No NMVr<br>(n=1004) | Risk difference | Relative Risk<br>Reduction | Odds ratio | P-value |
| --- | --- | --- | --- | --- | --- | --- |
| **Primary Outcome** |  |  |  |  |  |  |
| Broadly defined PASC | 273 | 347 | -0.074 (-0.114, -0.033) | 21.33% | 0.707 (0.585, 0.855) | <0.001 |
| Narrowly defined PASC | 221 | 278 | -0.057 (-0.094, -0.019) | 20.50% | 0.737 (0.601, 0.903) | 0.003 |
| **Secondary Outcomes** |  |  |  |  |  |  |
| **Individual Symptoms** |  |  |  |  |  |  |
| Constitutional symptoms | 108 | 128 | -0.018 (-0.046, 0.010) | 15.63% | 0.840 (0.639, 1.104) | 0.211 |
| CV and respiratory symptoms | 130 | 171 | -0.041 (-0.072, -0.010) | 23.98% | 0.725 (0.566, 0.928) | 0.010 |
| Gastrointestinal symptoms | 103 | 151 | -0.048 (-0.077, -0.019) | 31.79% | 0.646 (0.494, 0.844) | 0.001 |
| Nervous system and MSK symptoms | 46 | 54 | -0.008 (-0.027, 0.011) | 14.81% | 0.845 (0.564, 1.264) | 0.412 |
| Anxiety/mood disorder related symptoms | 157 | 211 | -0.054 (-0.088, -0.020) | 25.59% | 0.697 (0.555, 0.875) | 0.002 |
| **Healthcare Utilization** |  |  |  |  |  |  |
| Radiology diagnostic tests | 151 | 203 | -0.052 (-0.085, -0.019) | 25.62% | 0.698 (0.554, 0.881) | 0.002 |
| CV tests<br>(Echocardiogram and ambulatory rhythm monitors) | 20 | 38 | -0.018 (-0.033, -0.003) | 47.37% | 0.517 (0.298, 0.894) | 0.016 |
| AMB/Virtual Visit | 574 | 622 | -0.048 (-0.091, -0.005) | 7.72% | 0.820 (0.686, 0.980) | 0.029 |

Abbreviations: ER – Emergency Room, GI – Gastrointestinal

### Main Outcome

#### Between 30 to 180 days post-infection

For broadly defined PASC-associated symptoms, vaccinated patients treated with NMV-r within five□days of developing Covid-19 had lower odds of developing persistent or new PASC-associated symptoms. For example, PASC-associated symptoms occurred in 425 patients (42%) in the NMV-r cohort, as compared to 480 patients (48%) in the control cohort (OR 0.8 CI 0.67-0.96; p=0.01) between 30 to 180 days (OR=0.8, 95% CI 0.67-0.96, p=0.01). Similarly, for a narrow definition of PASC to three□symptom cluster, the use of NMV-r within five□days of development of Covid-19 was associated with lower odds of development of PASC-associated symptoms, which was recorded in 337 (34%) patients in the NMV-r and 404 (40%) of patients in the control cohort between 30 and 180 days (OR=0.75, CI 0.62-0.9, p=0.002).

#### Between 90 to 180 days post-infection

Even upon modifying the timeline for the development of new or persistent PASC-associated symptoms between 90 to 180 days, patients treated with NMV-r within five□days of diagnosis had lower odds of having persistent or new PASC-associated symptoms. Broadly defined PASC-associated symptoms occurred in 273 patients (27%) in the NMV-r cohort, as compared to 347 patients (35%) in the control cohort (OR 0.707, CI 0.59-0.86; p<0.001) from 90 to 180 days. In addition, narrowly defined PASC-associated symptoms based on three□symptom cluster was reported in 221 (22%) in the NMV-r cohort as compared to 278 (28%) patients in the control cohort (OR=0.7, CI 0.63-0.9, p=0.003).

### Other Outcomes

#### Between 30 to 180 days post-infection

Patients who had been treated with NMV-r within five□days of infection with Covid-19 had lower odds of having cardiovascular and respiratory symptoms (OR=0.72, CI 0.59-0.88, p=0.001); gastrointestinal symptoms (OR= 0.79, CI 0.63-0.98, p=0.03); and lower anxiety/mood disorders (OR=0.77, CI 0.63-0.94, p=0.01) between 30 to180 days post-infection.□There was no difference in constitutional and musculoskeletal, and nervous system symptoms. Cardiovascular testing was performed less frequently among those treated with NMV-r. However, there was no significant difference in the use of radiology diagnostic tests or ambulatory/virtual visits.

#### Between 90 to 180 days post-infection

Patients who had been treated with NMV-r within five□days of infection with Covid-19 had lower odds of having cardiovascular and respiratory symptoms (OR=0.72, CI 0.57-0.93, p=0.01); gastrointestinal symptoms (OR= 0.65, CI 0.49-0.84, p=0.001); anxiety/mood disorders (OR=0.69, 95% CI 0.55-0.88, p=0.002) and lower healthcare utilization including radiology and cardiovascular diagnostic tests and ambulatory/virtual visits between 90- and 180-days post-infection.

## Discussion

### Key Results

Our exploratory retrospective study of patients from a real-world population developing Covid-19 showed that use of NMV-r within five days of Covid-19 diagnosis was associated with 11 to 21% relative risk reduction for developing PASC-associated symptoms. A strength of this analysis is that it included only vaccinated patients who contracted Covid-19 when the Omicron variant was widely circulating, making it reflective of the current status of both the population’s high level of vaccine-or infection-induced immunity and the potential for Omicron to cause less severe disease than prior variants. This relative decrease in reported symptoms with treatment was observed using broad and narrow definitions of PASC symptoms between 30 to 180 days and 90 to 180 days. Cardiovascular, respiratory, gastrointestinal, and mood/cognitive disorder-related symptoms were recorded less frequently among those prescribed NMV-r than those not treated with NMV-r. No significant difference was noted for constitutional, musculoskeletal, and nervous system-related symptoms. Overall healthcare utilization, including radiology and cardiovascular diagnostic tests and ambulatory/virtual visits, was lower among those who received NMV-r.

Both quantitative and qualitative studies have demonstrated the complexity of diagnosing PASC in clinical settings. In addition, there are discrepancies in the definition of PASC, even among the National Institute of Health’s Researching COVID to Enhance Recovery (RECOVER) Initiative and the World Health Organization [11-13]. Therefore, to overcome some of the existing variability, we emphasize that our study does not explicitly address the diagnosis of PASC but uses the reporting of PASC-associated symptoms as an outcome. We further evaluated the reporting of these symptoms using two separate timelines 1) beyond the first 30 days and 2) beyond the first 90 days after the index diagnosis of Covid-19.

The pathophysiology of PASC remains uncertain, and no drugs have proven beneficial in its prevention or treatment. There is some correlation between initial disease severity and the likelihood of developing PASC, though even some people with mild Covid-19 can experience prolonged and disabling symptoms [3, 14]. There is evidence that vaccination reduces the incidence of PASC by 15-50% [15], and the incidence also appears to be lower with Omicron compared with infection with earlier variants [16]. Both these observations could result from reduced disease severity in people vaccinated and/or acquiring Omicron, a reduced severity explained by immunologic and viral factors. A small study demonstrated that patients diagnosed with PASC had evidence of persistent SARS-CoV-2 antigen detected in blood samples [14], raising the possibility of a future diagnostic test confirming PASC and potential interventions that reduce viral replication or the duration of acute disease will reduce the risk of PASC. A study of NMV-r treatment in people with PASC is ongoing [17].

Our results are similar to those reported in the study by the Department of Veterans Affairs [18]. However, in contrast with that study, our definition of PASC-associated symptoms is more diverse. Our patient population includes only those vaccinated, a much higher proportion of women (64%), and a longer follow-up period. The inclusion of more women in our study is vital, as PASC appears to be more common in women. The reason for this difference in incidence based on sex is unknown but has also been reported in diverse autoimmune conditions, raising the possibility of autoimmunity or immune dysregulation playing a vital role in the pathogenesis of PASC.

### Limitations

It is important to emphasize that, despite these observed favorable associations between NMV-r and reduced long-term symptoms, these data cannot prove that antiviral treatment reduces the incidence of PASC. As in many observational studies, the group receiving treatment differed significantly from those not receiving treatment, differences that could influence the results independent of the treatment they received. These biases could increase the risk of PASC in those accessing treatment and, conversely, decrease it. More severe symptoms could trigger reaching out for treatment and subsequent healthcare utilization, making PASC more likely in those receiving treatment. By contrast, patients who initially did not seek treatment but then later progressed may have fallen out of the 5 days from symptom onset treatment eligibility, rendering them unable to get NMV-r. While we controlled for differing baseline characteristics using propensity matching, residual confounding remains a significant potential limitation.

Several additional limitations intrinsic to the study design and the condition deserve mention. The retrospective design relied on the accuracy of the data entered in the EHR by point-of-care clinical providers; hence despite separately analyzing data for both broad and narrow-based PASC-associated symptoms, our definition may have lacked precision and may have been too inclusive. An intrinsic challenge in the study of PASC is the highly variable nature of the condition, which may involve multiple organ systems and a vast range of clinical severity, from mild fatigue to complete disability and no established diagnostic test or tests. Evidence for the condition’s heterogeneity is clear from the broad ranges reported for its incidence after Covid-19, from 6 to 54% [3],[4].

Limitations notwithstanding, our study supports the hypothesis that treating the acute infection with NMV-r may reduce the symptoms associated with PASC. It furthermore should reassure prescribers that treatment does not *increase* this risk; a potential concern given the widely observed viral and symptomatic rebound phenomenon. While a more accurate assessment of the effect of NMV-r treatment on the incidence of PASC would optimally come from the original placebo-controlled trials, EPIC-HR [5] and EPIC-SR [19], these analyses have not yet been conducted or reported. Studies of long-term outcomes are planned in more recent randomized trials, including COVID-OUT [20], ACTIV-6 [21], and PANORAMIC [22].

### Interpretation

In a retrospective analysis looking at long-term outcomes after diagnosis of Covid-19, these data show an association between treatment with NMV-r and a reduced incidence of the symptoms commonly reported with PASC. As the case severity of Covid-19 is much lower than when the pandemic first started, outcomes aside from hospitalization and death must be rigorously analyzed. As such, prospective studies of the potential effect of antiviral therapy on the incidence of PASC are urgently needed.□But, pending these data, our study does support the use of NMV-r to decrease even potential long-term complications of Covid-19, an important observation given the continued low rate of prescribing for this medication.

## Data Availability

All data produced in the present study are available upon request to the authors and is also readily available at https://trinetx.com/about-trinetx/community/.

## Acknowledgments

None

